# Health status and care utilization among Afghan refugees newly resettled in Calgary, Canada between 2011-2020

**DOI:** 10.1101/2024.06.21.24309182

**Authors:** Hannah Smati, Nour Hassan, Mohammad Yasir Essar, Fawzia Abdaly, Shayesta Noori, Rabina Grewal, Eric Norrie, Rachel Talavlikar, Julia Bietz, Sarah L. Kimball, Annalee Coakley, Avik Chatterjee, Gabriel E. Fabreau

## Abstract

**Background:** The United States and Canada have resettled over 120,000 Afghan refugees since August 2021, but sociodemographic and health status data remains sparse with investigations often limited to refugee entrance exams, standardized health screenings, or acute health settings.

**Methods:** This retrospective community-engaged cohort study investigated Afghan patients who received care between January 1, 2011 and December 31, 2020 at an interdisciplinary specialized refugee clinic in Calgary, Canada that provides care to newly arrived refugees. Two reviewers independently extracted and manually verified sociodemographic factors, medical diagnoses, and clinic utilization variables from patients’ electronic medical records, then coded patient diagnoses into ICD-10 codes and chapter groups. Diagnosis frequencies were calculated and stratified by age group and sex. We corroborated these findings with Afghan refugee co-investigators.

**Findings:** Among 402 Afghan refugee patients, 228 were adults (mean age 34·2 [SD 13] years), and 174 were children (mean age 7·5 [SD 5·4] years). We identified 1535 total individual diagnoses and classified them into 382 unique ICD-10 codes. Patients had a median 2 diagnoses each [IQR 0-6], 4 clinic visits across primary, specialty and multidisciplinary care annually, and an 11% appointment no-show rate. Among adults, the most frequent diagnoses were abdominal pain (26·3%, 60/228), mechanical back pain (20·2%, 46/228), and H. pylori infection (19·3%, 44/228). Among children, the most frequent diagnoses were upper respiratory tract infection (12·1%, 21/174), *Giardia* (10·3%, 18/174), and short stature (7·5%, 13/174).

**Interpretation:** Recently resettled Afghan refugees in Canada were relatively young, experienced diverse health characteristics, and had multi-specialty care engagement in their first two years after arrival. These findings may guide specialized healthcare provision to this inadequately characterized but growing population of refugee arrivals in North America and elsewhere.

**Funding:** Research grants from the M.S.I. Foundation and University of Calgary O’Brien Institute for Public Health

## Introduction

In August 2021, nearly 700,000 Afghans were forcibly displaced by intensified violence in Afghanistan amidst the United States’ military withdrawal and the Taliban’s government takeover.^1^ Before this crisis, nearly six million Afghans had been forcibly displaced over twenty years due to political conflict, violence, chronic poverty, and natural disasters, creating among the world’s largest protracted refugee crises.^2,3^ In response, the United States government launched “Operation Allies Welcome” in August 2021. Over the past two years, the Department of Homeland Security has resettled over 115,000 Afghan nationals in the United States via humanitarian parole or special immigrant visas.^4^ In parallel, the government of Canada has also committed to resettling Afghan refugees through special immigration and humanitarian pathways, so far resettling over 50,000 Afghan refugees since August 2021.^5^

Multiple studies have highlighted the complex health needs of resettled refugee populations that range from mental health issues to infectious disease to chronic non-communicable diseases.^6–10^ Despite North America’s growing Afghan refugee population, little clinical information exists about their health in the early resettlement period. A United States study that used medical examination data of incoming adult Afghan special immigrant visa holders found high rates of chronic disease such as obesity, hypertension, and tobacco use.^11^ Another study at a community health center in Toronto used a standardized trauma screen of adult Afghan refugees and found high rates of post-traumatic stress disorder (PTSD), associated closely with unemployment and inadequate social support.^12^

Most large cohort studies among Afghan refugees have been conducted in countries bordering Afghanistan or in European countries. Iran hosts the second largest Afghan refugee population globally. One cross-sectional study of Afghan refugee health referrals to Iran’s UNHCR offices found that the most frequent referrals were for ophthalmic disease, neoplasm, nephropathies, and ischemic heart disease.^13^ In Denmark, researchers found Afghan children experienced the lowest vaccination rates among all nationalities represented in a national asylee database.^14^ While large in scale, many of these investigations are limited to refugee entrance exams, standardized health screenings, or acute health settings, which may result in broad health characterizations that lack clinical granularity.

Recently arrived Afghan refugees’ unique health issues require further investigation with detailed, patient-level analysis to best assist domestic healthcare providers caring for them upon resettlement in North America. Thus, we conducted a detailed clinician and community-engaged investigation of the sociodemographic characteristics, health conditions, and clinic utilization among Afghan refugees at a large specialized interdisciplinary refugee medical home in Calgary, Canada between 2011 and 2020. This clinic has provided primary and multi-specialty care for over 12,000 refugees in Calgary since 2011 for up to two years after their arrival.^15^ Depending on their needs, patients receive care from family doctors, specialists, psychologists, dieticians, nurses, mental health therapists and other allied health professionals. We hypothesized that over a 10-year period, common age- and sex-stratified health trends could be identified that may inform North American healthcare providers caring for Afghan refugees who have arrived after August 2021.

## Methods

We manually reviewed electronic medical records among patients with at least one appointment at the Mosaic Refugee Health Clinic between January 1, 2011 and December 31, 2020 that had the text “Afghan” included in their profile, country of origin information, immigration documents, or those of their family members to ascertain Afghanistan was their country of origin. We also included newborn children of Afghan refugees born in Canada within the first 2 years of the mother’s arrival because they are an underrepresented population in research and may share social determinants of health with their refugee family members.^16–18^ Two reviewers independently reviewed electronically extracted data then manually verified all sociodemographic and utilization variables. We assessed total visits, primary care and specialty visits, multidisciplinary care team visits, missed visits, and duration of time people received care at the clinic. Internal medicine, pediatrics and obstetrician physicians provide subspecialty care on a referral basis in Canada, so these visits were counted as specialty care. Reviewers also manually clinical diagnoses from medical diagnoses, problem lists, treatment data, and past medical histories, then coded them into ICD-10-CA (International Classification of Diseases and Related Health Problems, the 10th Revision, Canada) codes using the online ICD-10-CA version 2016 tool.^19^ We grouped individual diagnostic codes into ICD-10 Chapters, an internationally standardized diagnostic nomenclature that organizes diagnoses generally by organ system.^19^ Given the short clinic follow up period and limitations of standardized coding diagnoses in capturing possible trauma-related health conditions, we compared our cohort’s ICD-10 diagnoses with a clinician consensus list of 18 ICD-10 diagnoses identified as suspected somatoform disorders among refugee patients that was generated during a recent analysis of trauma-related conditions among Yazidi patients.^20–22^ This list had been revised using a modified Delphi method for 3 rounds of voting and discussion to reach unanimous consensus between 5 clinician experts with experience in refugee clinical care.^22^

We summarized data using frequencies, means, standard deviation, median, quartiles, and range for continuous variables. We used World Health Organization age categories to stratify our data into four age groups (younger children aged 0-4 years, older children aged 5-11 years, adolescents aged 12-17 years and adults 18 years and older), and by sex among adults. Finally, all findings were further interpreted and validated by three Afghan refugee leader coinvestigators. We used STATA v.16 for all analyses (Stata Corp, College Station, Texas), and followed the STROBE reporting guidelines (checklist available in eMethods).^23^ This study received ethics approval and a consent waiver from the University of Calgary Conjoint Health Research Ethics Board (REB15-3264).

## Results

### Sociodemographic characteristics of patient sample

There were 402 Afghan refugee patients identified in our cohort. One hundred eighty-two (45·3%) patients were female, 228 (57·7%) were adults (mean age 34·2 years, SD 13 years), and 174 (43·3%) were children under 18 (mean age 7·5 years, SD 5·4 years). Their sociodemographic characteristics are presented in Table 1. Most patients came directly from Afghanistan (73·6%, 296/402 patients) and Pakistan (10·4%, 42/402), and had not stayed in a refugee camp prior to clinic presentation (89·3%, 349/402). Twenty-three patients (5·7%) were born in Canada to refugee parents and aged between 0·0-1·1 years at the time of our evaluation (mean 0·1 years, SD 0·2 years). Patients’ primary languages were Dari/Farsi (54·2%, 218/402) and Pashto (17·7%, 71/402). While most patients spoke no English (64·7%, 260/402), 24·4% (98/402) reported speaking English well. Among adults, 60·5% (138/228) were married and 65·8% (150/228) had children. There were 117 total families in our cohort (range 1-9 family members) and 3·4 members/family on average. Of the 347 patients who had documented refugee status, 47·0% (163/347) were government-sponsored refugees, or individuals resettled in Canada by the United Nations Refugee Agency or another humanitarian organization, while 40·1% (139/347) were privately sponsored by an individual already living in Canada. Asylum seekers, or individuals who apply for refugee protection upon or after arrival to Canada, made up 6·6% (23/347) of the overall cohort. The remaining 22 patients (6·3%) were citizens born in Canada.

**Table 1.** Sociodemographic characteristics among the Afghan refugee cohort by age group.

| Characteristics | Total<br>(n=402)<br>n (%) | Younger Children<br>0-4 years<br>(n=66)<br>n (%) | Older Children<br>5-11 years<br>(n=65)<br>n (%) | Adolescents<br>12-17 years<br>(n=43)<br>n (%) | Female adults<br>≥ 18 years<br>(n=112)<br>n (%) | Male Adults<br>≥ 18 years<br>(n=116)<br>n (%) |
| --- | --- | --- | --- | --- | --- | --- |
| **Gender** |  |  |  |  |  |  |
| Female | 182 (45.3) | 24 (36.4) | 24 (36.9) | 22 (51.2) | - | - |
| Male | 220 (54.7) | 42 (63.6) | 41 (63.1) | 21 (48.8) | - | - |
| **Previous Country of Residence** |  |  |  |  |  |  |
| Afghanistan | 296 (73.6) | 24 (36.4) | 33 (50.8) | 25 (58.1) | 102 (91.1) | 112 (96.6) |
| Pakistan | 42 (10.5) | 7 (10.6) | 20 (30.8) | 11 (25.6) | 2 (1.8) | 2 (1.7) |
| Canada <sup>a</sup> | 23 (5.7) | 23 (34.8) | 0 (0) | 0 (0) | 0 (0) | 0 (0) |
| India | 11 (2.7) | 3 (4.5) | 3 (4.6) | 2 (4.7) | 3 (2.7) | 0 (0) |
| Russia | 7 (1.7) | 3 (4.5) | 4 (6.2) | 0 (0) | 0 (0) | 0 (0) |
| Azerbaijan | 5 (1.2) | 3 (4.5) | 2 (3.1) | 0 (0) | 0 (0) | 0 (0) |
| Uzbekistan | 5 (1.2) | 1 (1.5) | 1 (1.5) | 1 (2.3) | 1 (0.9) | 1 (0.9) |
| Other <sup>b</sup> | 13 (3.2) | 2 (3.0) | 2 (3.1) | 4 (9.3) | 4 (3.6) | 1 (0.9) |
| **Refugee Category** |  |  |  |  |  |  |
| PSR | 139 (34.6) | 12 (18.2) | 25 (38.5) | 19 (44.2) | 46 (41.0) | 37 (31.9) |
| GAR | 163 (40.5) | 24 (36.4) | 33 (50.8) | 16 (37.2) | 43 (38.4) | 47 (40.5) |
| Canadian | 22 (5.5) | 22 (33.3) | 0 (0) | 0 (0) | 0 (0) | 0 (0) |
| Asylum seeker | 23 (5.7) | 2 (3.0) | 3 (4.6) | 0 (0) | 7 (6.3) | 11 (9.5) |
| Unknown | 55 (13.7) | 6 (9.1) | 4 (6.2) | 8 (18.6) | 16 (14.3) | 21 (18.1) |
| **Primary Language** |  |  |  |  |  |  |
| Dari/Farsi | 218 (54.2) | 9 (13.6) | 39 (60) | 25 (58.1) | 74 (66.1) | 71 (61.2) |
| Pashto | 71 (17.7) | 13 (19.7) | 9 (13.8) | 7 (16.3) | 19 (17) | 23 (19.8) |
| Hindi | 33 (8.2) | 3 (4.5) | 10 (15.4) | 4 (9.3) | 9 (8) | 7 (6) |
| N/A | 33 (8.2) | 32 (48.5) | 0 (0) | 1 (2.3) | 0 (0) | 0 (0) |
| English | 21 (5.2) | 5 (7.6) | 4 (6.2) | 4 (9.3) | 2 (1.8) | 6 (5.2) |
| Unknown | 9 (2.2) | 0 (0) | 1 (1.5) | 0 (0) | 75 (67) | 80 (69) |
| Multani | 5 (1.2) | 1 (1.5) | 1 (1.5) | 0 (0) | 2 (1.8) | 1 (0.9) |
| Other <sup>c</sup> | 12 (2.9) | 3 (4.5) | 1 (1.5) | 2 (4.6) | 4 (3.6) | 2 (1.8) |
| English Proficiency |  |  |  |  |  |  |
| No English | 260 (64.7) | 59 (89.4) | 50 (76.9) | 25 (58.1) | 78 (69.6) | 48 (41.4) |
| Some English | 44 (10.9) | 1 (1.5) | 8 (12.3) | 7 (16.3) | 14 (12.5) | 14 (12.1) |
| Speaks English Well | 98 (24.4) | 6 (9.1) | 7 (10.8) | 11 (25.6) | 20 (17.9) | 54 (46.6) |
| Marital Status |  |  |  |  |  |  |
| Single | 244 (60.7) | 66 (100) | 65 (100) | 41 (95.3) | 31 (27.7) | 41 (35.3) |
| Married | 140 (34.8) | 0 (0) | 0 (0) | 2 (4.7) | 67 (59.8) | 71 (61.2) |
| Widowed | 15 (3.7) | 0 (0) | 0 (0) | 0 (0) | 14 (12.5) | 1 (0.9) |
| Number of children (n=228) <sup>d</sup> |  |  |  |  |  |  |
| Zero | 78 (34.2) | - | - | - | 29 (25.9) | 49 (42.2) |
| 1-2 | 59 (25.9) | - | - | - | 32 (28.6) | 27 (23.3) |
| 3-4 | 59 (25.9) | - | - | - | 34 (30.4) | 25 (21.6) |
| ≥ 5 | 32 (14.0) | - | - | - | 17 (15.2) | 15 (12.9) |
| Camp Exposure |  |  |  |  |  |  |
| No Camp Exposure | 359 (89.3) | 63 (95.5) | 53 (81.5) | 37 (86) | 100 (89.3) | 106 (91.4) |
| Camp Exposure | 43 (10.7) | 3 (4.5) | 12 (18.5) | 6 (14) | 12 (10.7) | 10 (8.6) |

*Abbreviations:* PSR, Privately-sponsored refugees (sponsored by an individual living in Canada to resettle in the same community as their sponsor); GAR, Government-Assisted Refugee (referred to Canada for resettlement by the United Nations Refugee Agency or another humanitarian organization).
<sup>a</sup> Patients with “Canada” as their country of residence were infants born in Canada post-arrival of the family.
<sup>b</sup> Other countries were Iran, England, Georgia, Indonesia, Iraq, Mexico, Tajikistan, and Ukraine.
<sup>c</sup> Other languages were Uzbek, Nooristan, Russian, Urdu, Arabic, and Punjabi.
<sup>d</sup> The proportions for number of children are reported for adults only

### Health characteristics

During the study period, we recorded 1535 individual diagnoses, classified them into 382 unique ICD-10 codes, and grouped into 22 ICD-10 chapters. Patients had a median of two diagnoses each [interquartile range 0-6]. One hundred four patients (25·9%) had no diagnoses identified in their records. Adult women had the most recorded diagnoses (6·3 diagnoses/patient) relative to adult men (4·2 diagnoses/patient) and children (2·0 diagnoses/patient). The most common individual diagnoses are shown in Figure 1. Among adults, the most frequent overall diagnoses were abdominal pain (26·3%, 60/228), mechanical back pain (20·2%, 46/228), and *H. pylori* infection (19·3%, 44/228). Among children, the most frequent diagnoses were upper respiratory tract infection (12·1%, 21/174), *Giardia* (10·3%, 18/174), and short stature (7·5%, 13/174) though frequency varied considerably between age groups. For younger children aged 0-5 years old, the most coded diagnoses included upper respiratory tract infection (28·8%, 19/66), short stature (9·1%, 6/66), constipation (9·1%, 6/66), and eczema (9·1%, 6/66). For adolescents, the most frequent diagnoses were *H. Pylori* infection (16·3%, 7/43), acne (14·0%, 6/43), and *Giardia* (11·6%, 5/43 patients). The complete list of diagnoses and frequency by age group are shown in eTables 1-5.

**Figure 1.**
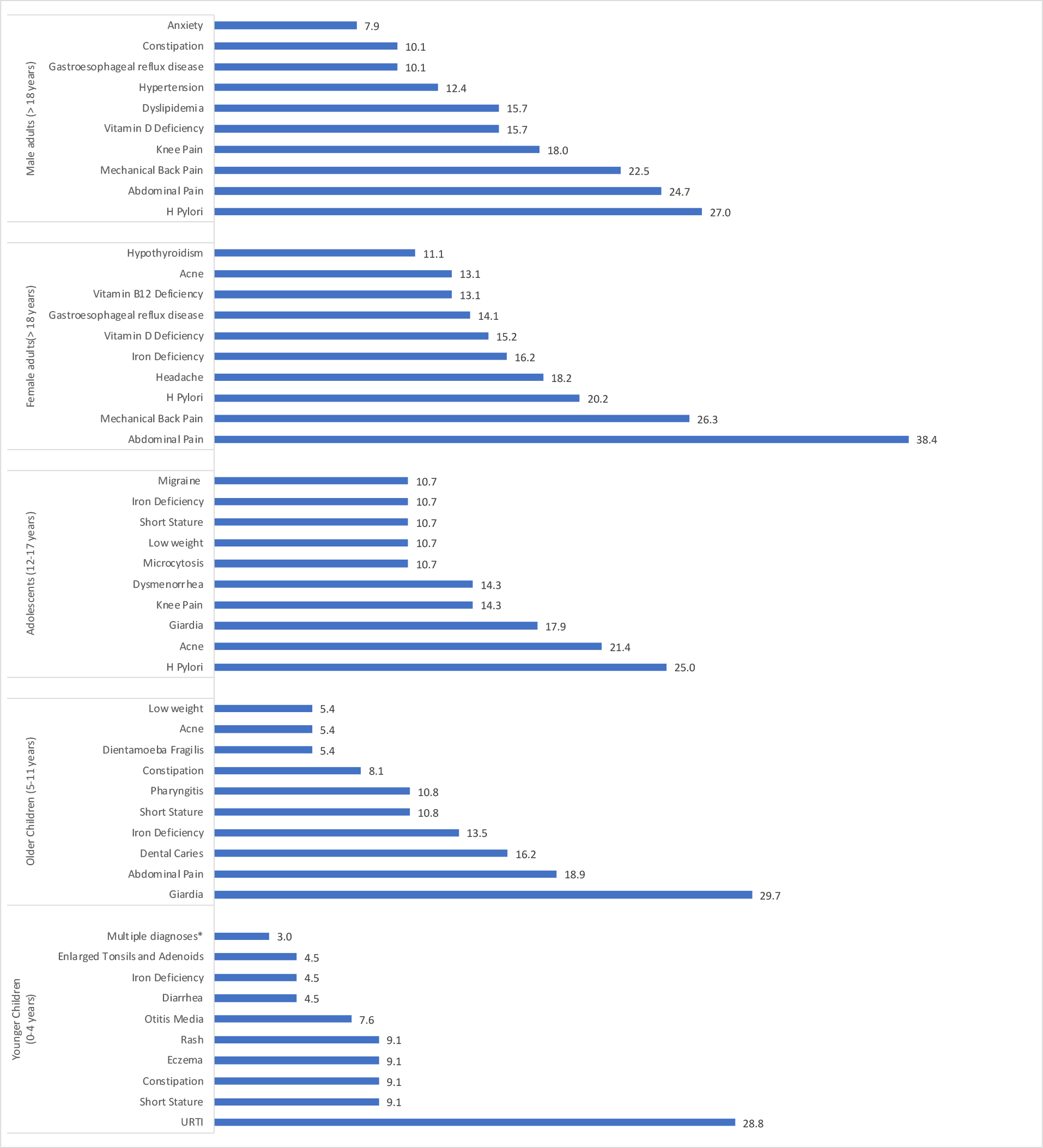
Ten most prevalent ICD-10 diagnoses by age group (Prevalence, %). *There were multiple diagnoses of equal prevalence (3.0%, 2/10) as follows: Giardia, Skin lesions, Iron Deficiency Anemia, Beta Thalassemia Trait, Strabismus, Pharyngitis, Pain in Limb, Phimosis, Balanitis, Sacral Dimple, Low Weight, Abnormal Lead Level in Blood

Table 2 shows the most common ICD-10 chapters among adults. After non-specific symptoms (46·1%, 105/228), most adult patients had musculoskeletal/connective tissue disorders (39·0%, 89/228), endocrine, nutritional, and metabolic disorders (37·7%, 86/228), and infections (35·5%, 81/228). The most frequent musculoskeletal or connective tissue related diagnoses were mechanical back pain (20·2%, 46/228), knee pain (11·8%, 27/228 patients), and pain in limb (7·5%, 17/228). The most prevalent nutritional or metabolic related disorder was vitamin D deficiency (12·3%, 28/228), followed by dyslipidemia (10·1%, 23/228) and iron deficiency (8·3%, 19/228). Among infections, *H. pylori* and *Giardia* were the two most frequently diagnosed infections for both men and women, followed by bacterial vaginosis for women (8·9%, 10/112) and *Dientamoeba fragilis* for men (4·3%, 5/116). Gastrointestinal diseases were also frequently reported (33·5%, 63/228), chief among them gastroesophageal reflux disease (GERD) (27·6%, 23/228) and constipation (8·8%, 20/228). Mental and behavioral health disorders affected 18·9% of adults in the cohort, with depression (6·6%, 15/228), anxiety (4·8%, 11/228), and PTSD (4·8%, 11/228) being the most frequently coded.

**Table 2.** Top 10 most common ICD-10 chapters and their 5 common prevalent diagnoses among Afghan refugee adults (n = 228)

| Rank | ICD-10 chapter | ICD-10 code | ICD-10 description | n | Prevalence (%) |
| --- | --- | --- | --- | --- | --- |
| 1 | Chapter 18: Symptoms, signs and abnormal clinical findings | R10 | Abdominal and pelvic pain | 60 | 26.3 |
|  |  | R51 | Headache | 25 | 11.0 |
|  |  | R07 | Pain in throat and chest | 13 | 5.7 |
|  |  | R21 | Rash and other nonspecific skin eruption | 12 | 5.3 |
|  |  | R42 | Dizziness and giddiness | 9 | 3.9 |
| 2 | Chapter 4: Endocrine, nutritional, and metabolic diseases | E55 | Vitamin D Deficiency | 29 | 12.7 |
|  |  | E78 | Dyslipidemia | 23 | 10.1 |
|  |  | E61.1 | Iron deficiency | 19 | 8.3 |
|  |  | E53 | Vitamin B12 Deficiency | 16 | 7.0 |
|  |  | E03.9 | Hypothyroidism, unspecified | 13 | 5.7 |
| 3 | Chapter 12: Diseases of the skin and subcutaneous tissue | L70 | Acne | 18 | 7.9 |
|  |  | L65 | Other nonscarring hair loss | 11 | 4.8 |
|  |  | L30.9 | Dermatitis, unspecified | 7 | 3.1 |
|  |  | L81.1 | Chloasma | 5 | 2.2 |
|  |  | L85.3 | Xerosis cutis | 5 | 2.2 |
| 4 | Chapter 1: Certain infectious and parasitic diseases | B98.0 | Helicobacter pylori | 44 | 19.3 |
|  |  | A07.1 | Giardiasis | 17 | 7.5 |
|  |  | A07.8 | Other specified protozoal intestinal diseases | 10 | 4.4 |
|  |  | A15.9 | Respiratory tuberculosis, unspecified | 7 | 3.1 |
|  |  | B00.1 | Herpes simplex infections | 4 | 1.8 |
| 5 | Chapter 10: Diseases of the respiratory system | J06 | Acute upper respiratory infections of multiple and unspecified sites | 11 | 4.8 |
|  |  | J02.9 | Acute pharyngitis, unspecified | 8 | 3.5 |
|  |  | J32.9 | Chronic sinusitis, unspecified | 4 | 1.8 |
|  |  | J30 | Vasomotor and allergic rhinitis | 2 | 0.9 |
|  |  | J30.1 | Allergic rhinitis due to pollen | 2 | 0.9 |
| 6 | Diseases of the digestive system | K21 | Gastro-esophageal reflux disease | 23 | 10.1 |
|  |  | K59 | Constipation | 20 | 8.8 |
|  |  | K29.7 | Gastritis, unspecified | 13 | 5.7 |
|  |  | K02 | Dental caries | 10 | 4.4 |
|  |  | K64 | Hemorrhoids and perianal venous thrombosis | 6 | 2.6 |
| 7 | Diseases of the musculoskeletal system and connective tissue | M54.5 | Low back pain | 46 | 20.2 |
|  |  | M25.5 | Pain in joint | 27 | 11.8 |
|  |  | M79.6 | Pain in limb | 17 | 7.5 |
|  |  | M19 | Other arthrosis | 8 | 3.5 |
|  |  | M72.2 | Plantar fascial fibromatosis | 4 | 1.8 |
| 8 | Diseases of the blood and blood-forming organs and certain disorders involving the immune mechanism | D50 | Iron deficiency anemia | 12 | 5.3 |
|  |  | D64 | Other anemias | 8 | 3.5 |
|  |  | D50.8 | Microcytosis | 7 | 3.1 |
|  |  | D56.3 | Thalassemia trait | 7 | 3.1 |
|  |  | D70 | Agranulocytosis | 5 | 2.2 |
| 9 | Chapter 8: Diseases of the ear and mastoid process | H91 | Other hearing loss | 8 | 3.5 |
|  |  | H61.2 | Impacted cerumen | 6 | 2.6 |
|  |  | H72 | Perforation of tympanic membrane | 3 | 1.3 |
|  |  | H92.0 | Otalgia | 2 | 0.9 |
|  |  | H60.9 | Otitis externa, unspecified | 1 | 0.4 |
| 10 | Chapter 14: Diseases of the genitourinary system | N76.0 | Acute vaginitis | 10 | 4.4 |
|  |  | N94.6 | Dysmenorrhea, unspecified | 8 | 3.5 |
|  |  | N39.0 | Urinary tract infection, site not specified | 6 | 2.6 |
|  |  | N89.8 | Other specified noninflammatory disorders of vagina | 6 | 2.6 |
|  |  | N94.1 | Dyspareunia | 6 | 2.6 |

Table 3 shows the most common ICD-10 chapters among children. Like adults, non-specific symptoms and signs were the most commonly reported conditions (21·8%, 38/174) among pediatric patients and included abdominal pain (5·2%, 9/174), rash (4·0%, 7/174), and low weight (4·0%, 7/174). Growth and nutritional deficiencies were also commonly reported, with 6·3% across all age groups diagnosed with iron deficiency (11/174) and 7·5% diagnosed with short stature (13/174). Lastly, infections were also prevalent among children (20·1%, 35/174). The most frequently diagnosed infections were *Giardia* (10·3%, 18/174) and *H. pylori* (4·02%, 7/174). Sixty-one percent of patients with *Giardia* were aged 5-11 years (11/18), while only adolescents were diagnosed with *H. pylori* infections (7/7).

**Table 3.**
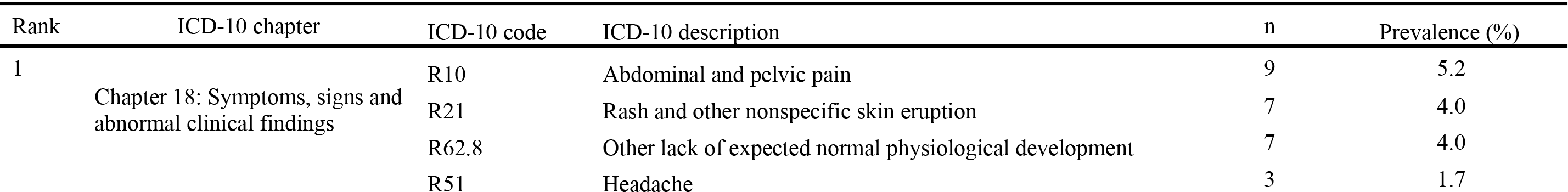
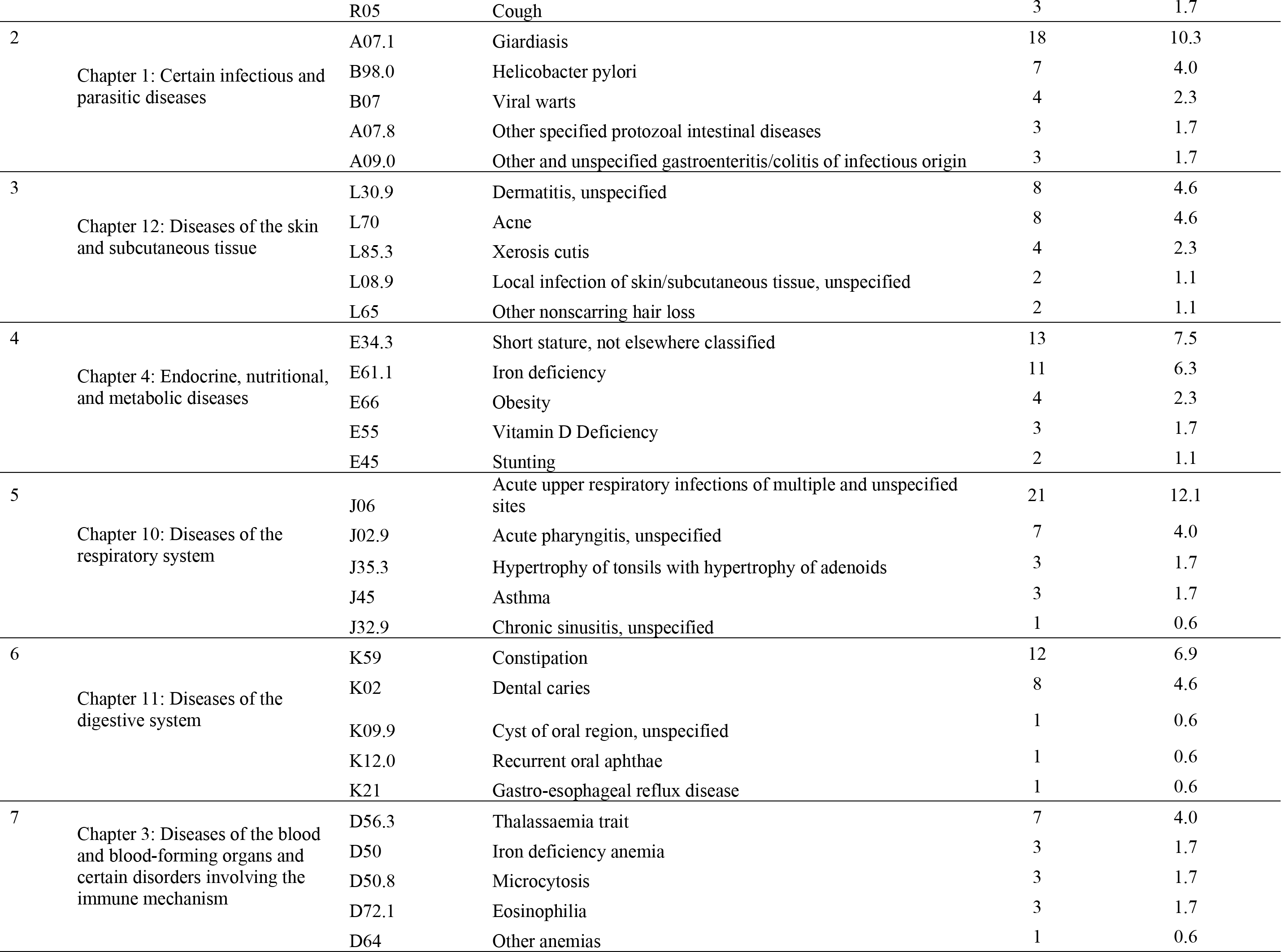
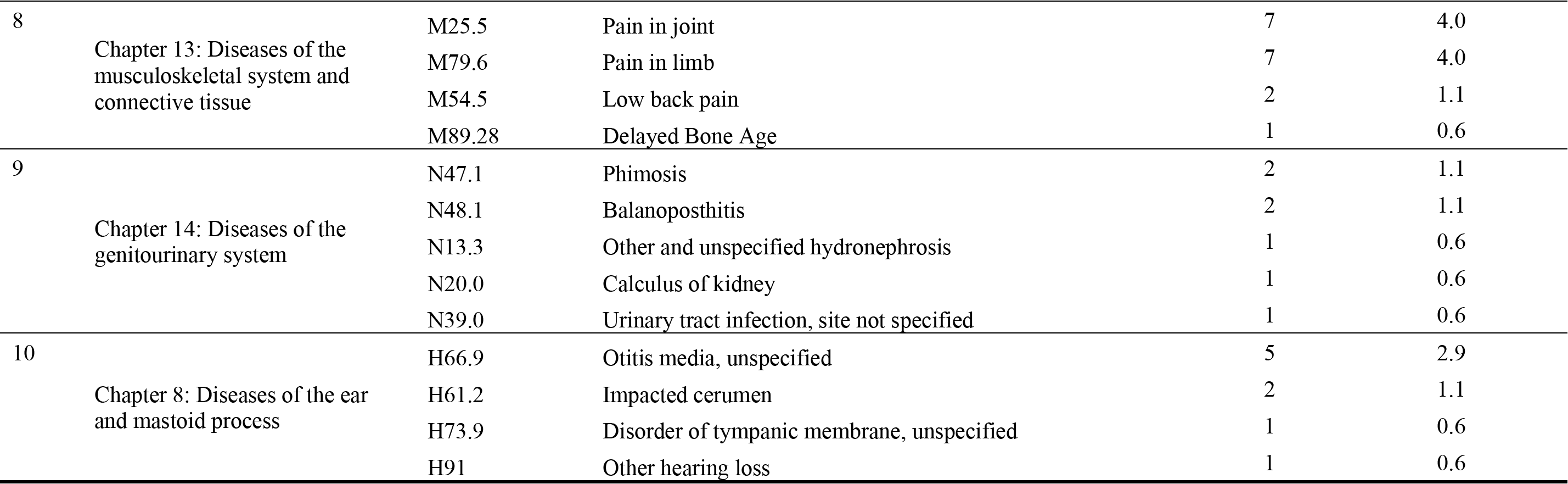
Top 10 most common ICD-10 chapters and their 5 common prevalent diagnoses among Afghan refugee children (n = 174)

Overall, 18% of total diagnoses coded among our cohort (285/1535 diagnoses) were physical symptoms recently identified as suspected somatoform disorders by refugee clinician consensus.^22^ Table 4 shows the prevalence of ICD-10 diagnoses identified as clinician consensus-derived suspected somatoform disorders by age group. The most frequently coded diagnosis suspicious for a somatoform disorder by clinicians for both adults and children was abdominal and pelvic pain (26·3%, 60/228 adults; 6·32%, 11/174 pediatric patients), followed by low back pain for adults (20·2%, 46/228) and joint pain for children (4·0%, 7/174).

**Table 4.** Prevalence of ICD-10 diagnoses identified as clinician consensus-derived suspected somatoform disorders among Afghan refugees (n = 402)

| ICD-10 code | ICD-10 description | Total<br>(n=402)<br>n (%) | Children<br>0-11 years<br>(n=66)<br>n (%) | Adolescents<br>12-17 years<br>(n=43)<br>n (%) | Female adults<br>≥ 18 years<br>(n=112)<br>n (%) | Male Adults<br>≥ 18 years<br>(n=116)<br>n (%) |
| --- | --- | --- | --- | --- | --- | --- |
| F80.9 | Developmental disorder of speech and language, unspecified | 1 (0.2) | 1 (0.8) | 0 (0) | 0 (0) | 0 (0) |
| F98 | Enuresis | 0 (0) | 0 (0) | 0 (0) | 0 (0) | 0 (0) |
| G43 | Migraine | 17 (4.2) | 1 (0.8) | 3 (7.0) | 11 (9.8) | 2 (1.7) |
| G44.2 | Tension-type headaches | 9 (2.2) | 1 (0.8) | 0 (0) | 8 (7.1) | 0 (0) |
| K58 | Irritable bowel syndrome | 3 (0.7) | 0 (0) | 0 (0) | 1 (0.9) | 2 (1.7) |
| L65 | Hair loss | 13 (3.2) | 0 (0) | 2 (4.7) | 6 (5.4) | 5 (4.3) |
| M25.5 | Joint pain | 35 (8.7) | 3 (2.3) | 4 (9.3) | 12 (10.7) | 16 (13.8) |
| M54.5 | Low back pain | 48 (11.9) | 0 (0) | 2 (4.7) | 26 (23.2) | 20 (17.2) |
| N76.0 | Acute vaginitis | 10 (2.5) | 0 (0) | 0 (0) | 10 (8.9) | 0 (0) |
| N89.8 | Vaginal discharge | 6 (1.5) | 0 (0) | 0 (0) | 6 (5.4) | 0 (0) |
| R07 | Chest pain | 13 (3.2) | 0 (0) | 0 (0) | 6 (5.4) | 7 (6.0) |
| R10 | Abdominal and pelvic pain | 71 (17.7) | 8 (6.1) | 3 (7.0) | 38 (33.9) | 22 (19.0) |
| R20.8 | Burning sensation | 2 (0.5) | 0 (0) | 0 (0) | 1 (0.9) | 1 (0.9) |
| R42 | Dizziness | 9 (2.2) | 0 (0) | 0 (0) | 5 (4.5) | 4 (3.4) |
| R47.8 | Speech disturbances, unspecified | 0 (0) | 0 (0) | 0 (0) | 0 (0) | 0 (0) |
| R51 | Headaches | 28 (7.0) | 0 (0) | 3 (7.0) | 18 (16.1) | 7 (6.0) |
| R52 | Chronic pain | 9 (2.2) | 0 (0) | 0 (0) | 5 (4.5) | 4 (3.4) |
| R53 | Fatigue | 11 (2.7) | 0 (0) | 1 (2.3) | 7 (6.3) | 3 (2.6) |

### Clinic utilization

On average, patients had a median of four clinic visits annually [interquartile range 2-7 visits] and spent an average of 12·4 [5·3-21·4] months at the refugee clinic before transitioning to a community-based clinic (Table 5). Of the total visits, 66·3% (3000/4526) were with a primary care physician, 26·6% (1202/4526) were with a multi-disciplinary team member, and 7·2% (325/4526) were with a specialist at the clinic. Visit types are summarized in Figure 2. Most specialty appointments were with psychiatry (35·4%, 115/325), followed by OB/GYN (16·9%, 55/325) and internal medicine (10·2%, 33/325). There was one pediatric specialty visit for an adult patient that was scheduled and billed as a pediatric appointment in which a pediatrician called a child’s parent to give an update. Within the multidisciplinary appointments, 30·9% (449/1202) were with a social worker, 14·5% (174/1202) with a health liaison who helps patients navigate healthcare and immigration systems, 10% (120/1202) with a psychologist, and 7% (85/1202) with a dietician. The overall appointment no-show rate for missed visits was 11·1% over the 10-year study period. We summarize the no-show rate by year in eTable 6.

**Figure 2.**
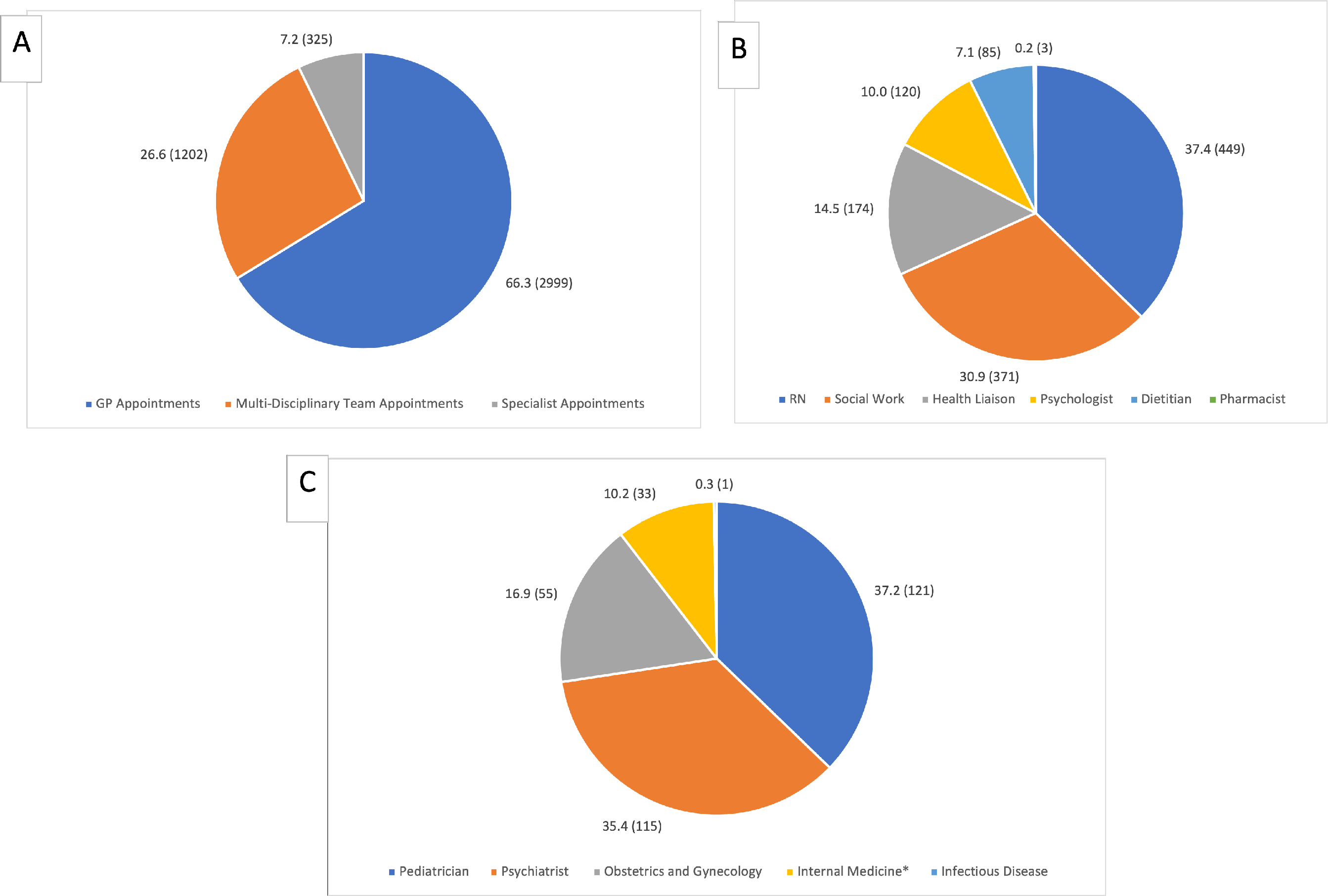
Clinic appointment type and frequency among the Afghan refugee cohort. A: Frequency of overall appointment types, percentage (number of clinic visits). B: Frequency of multi-disciplinary appointments, percentage (number of clinic visits). C: Frequency of specialty appointments, percentage (number of clinic visits). *In Canada, internal medicine physicians provide subspecialty care on a referral basis in Canada. Internal medicine visits were counted as specialty care.

**Table 5.** Average clinic utilization among the Afghan refugee cohort by age group.

| Clinic use measures | Total (N=402) | Younger Children<br>0-4 years<br>(n=66) | Older Children<br>5-11 years<br>(n=65) | Adolescents<br>12-17 years<br>(n=43) | Female adults<br>≥ 18 years<br>(n=112) | Male Adults<br>≥ 18 years<br>(n=116) |
| --- | --- | --- | --- | --- | --- | --- |
| Median number of months in clinic<br>[Interquartile Range] | 12.4 [5.3-21.4] | 13.5 [5.5-22.1] | 11.4 [5.1-21.4] | 11.3 [5.8-19.2] | 14.9 [5.8-24.3] | 13.0 [3.4-20.9] |
| Median appointments/person/year<br>[Interquartile Range] | 4 [2-7] | 4 [2-6] | 3 [1-4] | 3 [1-5] | 6 [3-11] | 4 [2-7] |

## Discussion

We present retrospective longitudinal, granular health and clinic utilization information about an inadequately characterized yet rapidly growing Afghan refugee population in Canada that may help guide healthcare for ongoing refugee arrivals in North America. We found that Afghan refugees resettled over the preceding decade primarily arrived in families and were predominantly Dari/Farsi-speaking, with nearly half of adult males proficient in English, potentially suggesting a cohort of interpreters. Afghan refugees in our cohort have diverse health needs with frequent clinic use and specialty consultation in the early resettlement period, particularly among adult women who had higher proportions of diagnoses and clinic utilization. Our study identified health conditions commonly encountered in refugee populations but also in general primary care practice. Consistent with previous research, we found key diagnoses among adult refugees were nutritional deficiencies and gastrointestinal infections.^11,24^ However, we also found that chronic diseases such as hypertension, dyslipidemia, and GERD were prevalent, particularly among male adults. This finding corroborates a cross-sectional study of Afghan refugees in Iran that found high prevalence of dyslipidemia and hypertension in addition to other non-communicable disease.^25^ Among refugee children, we similarly found malnutrition and low acuity conditions commonly encountered in the general pediatric population, but also less common infectious diseases such as intestinal parasites. These findings are consistent with published reports of endemic intestinal parasites among newly arrived pediatric refugees in the U.S., and support North American refugee health guidelines on screening for intestinal parasites.^26–28^

Unlike other analyses of post-resettlement Afghan refugees, our cohort had a lower observed prevalence of diagnosed mental illnesses like PTSD relative to other organ systems.^12,29^ We suspect that formal diagnoses of mental health disorder may have been limited by the short period that patients were followed in the refugee clinic, with generalists primarily reporting non-specific symptoms until other organic diagnoses could be ruled out or a formal psychiatric diagnosis could be made. In contrast, a study conducted in Turkey among Afghan refugees reported high rates of depression (50%) and anxiety (41%), indicating a higher prevalence of mental health conditions than observed in our cohort.^30^ Daily stressors such as financial security, language barrier, access to healthcare during migration journey as well as traumatic experiences are likely important contributing factors.^31^ Unlike other specialized refugee clinics, ours does not systematically screen new patients for mental health conditions in the early post-resettlement period, instead preferring first to build trust among clinicians and patients before screening. This differs from other refugee healthcare models that use validated refugee mental health screening tools such as the Refugee Health Screener-15 immediately upon arrival, and could represent an ascertainment bias that may account for the lower frequency of diagnosed mental illness observed in our cohort.^32^

Somatic symptoms associated with trauma are well documented among refugees, including abdominal and joint pain, but remain challenging to diagnose in electronic medical record systems.^20,21^ When compared with a recent Yazidi refugee analysis using expert clinician consensus to identify potential trauma-related conditions, we found that one-fifth of diagnoses (18%) made among our cohort were on a consensus list of non-specific diagnoses clinicians suspected represented somatoform disorders.^22^ This proportion suggest the importance of adopting a culturally sensitive approach to diagnosing mental health illnesses among Afghan and other refugees despite potential reluctance, at times, to openly discuss mental health.^33^

Following the Taliban’s takeover, our team has observed a significant linguistic and cultural shift among Afghan refugees settled locally. There has been a notable emergence of Pashto as the predominant language, alongside Dari, among new arrivals. This linguistic transition may complicate pre-existing interpretation and cultural brokering services already established in North America and present additional challenges for newly arrived Afghans to access healthcare and integrate socially for both Dari and Pashto speakers.

Despite the relatively short duration Afghan refugee patients spent at the refugee clinic before transitioning to the general healthcare system, we found many patients in the immediate post-resettlement period benefited from multidisciplinary primary care at the clinic such as social work, psychology, and nutrition, with relatively low no-show rates for a socially vulnerable population.^34–36^ Importantly, this detailed health assessment completed so early after arrival likely reflects the health conditions of millions of Afghan refugees displaced abroad. Notably, 10% of our cohort had resided in a refugee camp, and 14% had been displaced to a nearby Eastern Asian country nearby to Afghanistan prior to migration.

Our study had several limitations. First, it had a relatively small sample size of Afghan refugees resettled in Calgary at a single specialized refugee clinic, which may limit the generalizability of the findings among other Afghan refugees resettled elsewhere. Second, patients on average received care for a relatively short time in the clinic, so diagnoses noted by clinicians at the time of extraction may have been more symptom-based and not the ultimate diagnosis. Third, over a 10-year study period, the clinic’s screening practices, such as empirically treating for Vitamin D deficiency or universal screening with stool ova and parasites, have inevitably evolved and may skew the prevalence of certain diagnoses based on the year. Lastly, the study period does not include 2021 arrivals, and may not be generalizable to the most recent wave of Afghan arrivals. Challenges in resettlement process, such as language barriers, cultural differences, employment, and healthcare access may have impacted the healthcare utilization patterns among newly arrived Afghan refugees in ways that were not fully captured in the present study. Of note, the U.S.’s Operation Allies Welcome has prioritized interpreters and individuals who worked with military forces, a group which may have different socioeconomic status than previously resettled Afghan refugee cohorts and thus affect prevalence of nutritional deficiencies and undertreated chronic disease.^37,38^

As the United States and Canadian governments continue to rapidly resettle Afghan refugees in North America, a nuanced understanding of this vulnerable population’s health status and sociodemographic profile is critical for healthcare delivery. Despite its limitations, our study provides new, patient-level information about the health status and multi-specialty care engagement of a longitudinal cohort of Afghan refugees resettled in Canada early after arrival. Our findings may inform health providers caring for new Afghan arrivals, assist refugee healthcare model design for the early resettlement period, and inform local and global refugee health policy to support displaced Afghan populations in North America and elsewhere amid the current and growing refugee crisis.^2^

## Data Availability

All data produced in the present study are available upon reasonable request to the authors

## Acknowledgements

We would like to thank the healthcare providers, staff and especially the patients at the Mosaic Refugee Health Clinic for their many contributions. Our sincere thanks for support from the Refugee Health YYC research team and the institutional support provided by the O’Brien Institute for Public Health, at the University of Calgary Cumming School of Medicine.

## Data Access and Author Contributions

Authors NH, RG, EN and GF had full access to the data utilized in this study. Authors NH, RG and EN contributed to data collection and analysis. Authors HS, NH, RT, JB, EN, YE, FA, SN, AC, and GEF contributed to the design, draft, and revision of the study manuscript.

## Supplementary Materials

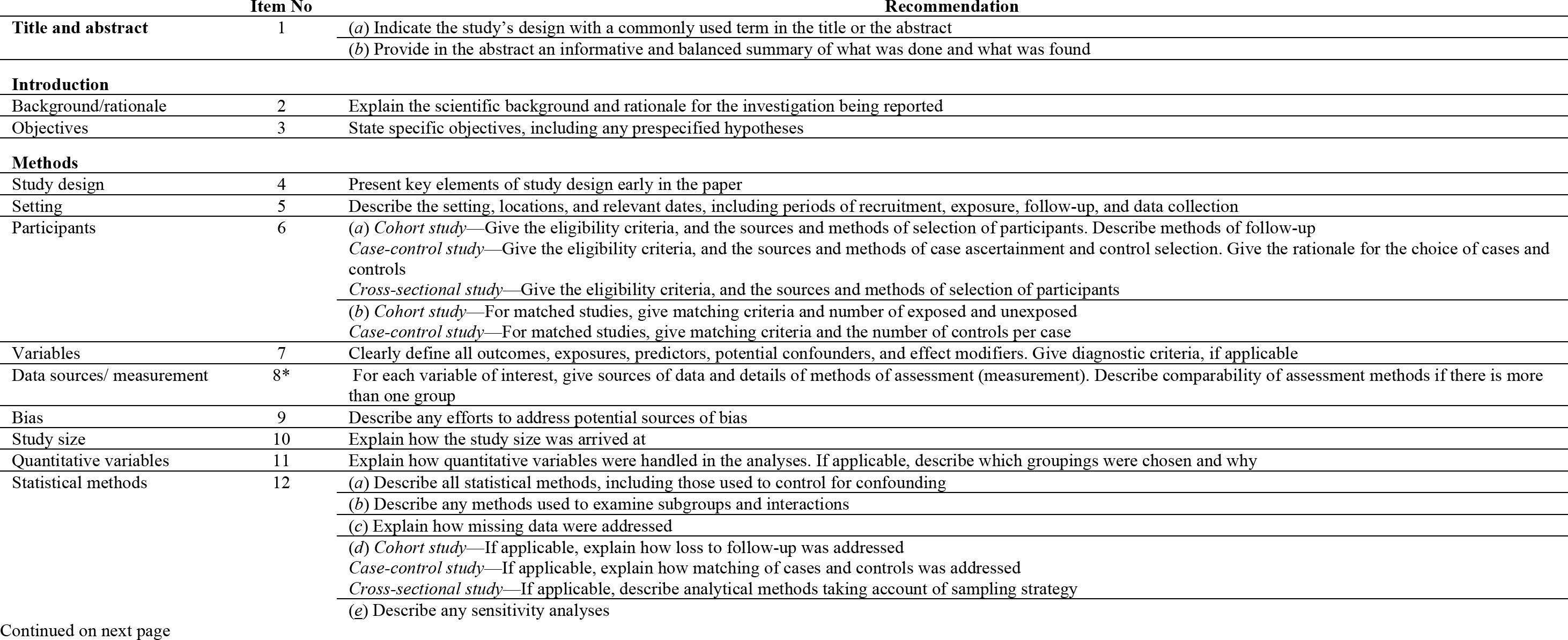
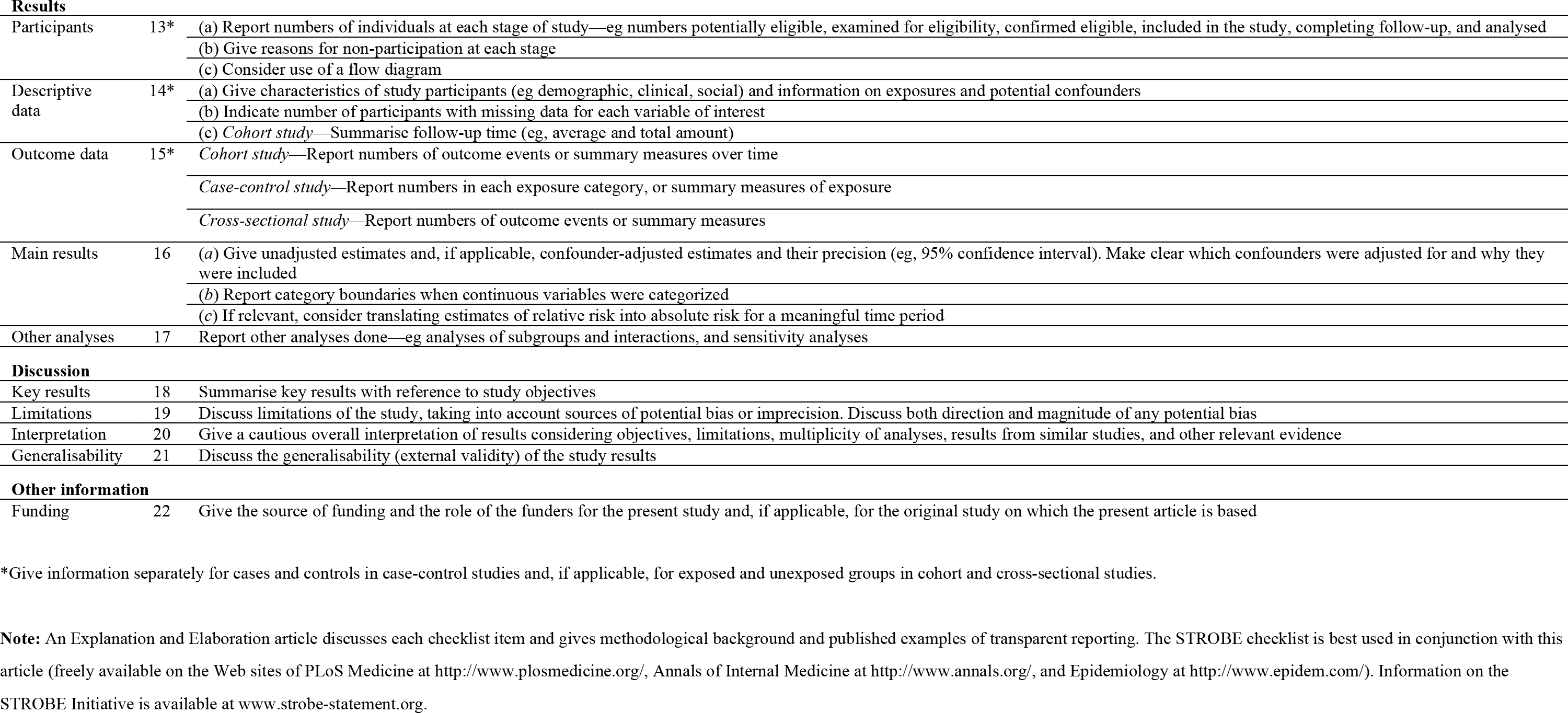
eMethods. STROBE Statement—checklist of items that should be included in reports of observational studies.

|  | Item No | Recommendation |
| --- | --- | --- |
| **Title and abstract** | 1 | (a) Indicate the study's design with a commonly used term in the title or the abstract<br>(b) Provide in the abstract an informative and balanced summary of what was done and what was found |
| **Introduction** |  |  |
| Background/rationale | 2 | Explain the scientific background and rationale for the investigation being reported |
| Objectives | 3 | State specific objectives, including any prespecified hypotheses |
| **Methods** |  |  |
| Study design | 4 | Present key elements of study design early in the paper |
| Setting | 5 | Describe the setting, locations, and relevant dates, including periods of recruitment, exposure, follow-up, and data collection |
| Participants | 6 | (a) *Cohort study*—Give the eligibility criteria, and the sources and methods of selection of participants. Describe methods of follow-up<br>*Case-control study*—Give the eligibility criteria, and the sources and methods of case ascertainment and control selection. Give the rationale for the choice of cases and controls<br>*Cross-sectional study*—Give the eligibility criteria, and the sources and methods of selection of participants<br>(b) *Cohort study*—For matched studies, give matching criteria and number of exposed and unexposed<br>*Case-control study*—For matched studies, give matching criteria and the number of controls per case |
| Variables | 7 | Clearly define all outcomes, exposures, predictors, potential confounders, and effect modifiers. Give diagnostic criteria, if applicable |
| Data sources/ measurement | 8* | For each variable of interest, give sources of data and details of methods of assessment (measurement). Describe comparability of assessment methods if there is more than one group |
| Bias | 9 | Describe any efforts to address potential sources of bias |
| Study size | 10 | Explain how the study size was arrived at |
| Quantitative variables | 11 | Explain how quantitative variables were handled in the analyses. If applicable, describe which groupings were chosen and why |
| Statistical methods | 12 | (a) Describe all statistical methods, including those used to control for confounding<br>(b) Describe any methods used to examine subgroups and interactions<br>(c) Explain how missing data were addressed<br>(d) *Cohort study*—If applicable, explain how loss to follow-up was addressed<br>*Case-control study*—If applicable, explain how matching of cases and controls was addressed<br>*Cross-sectional study*—If applicable, describe analytical methods taking account of sampling strategy<br>(e) Describe any sensitivity analyses |
| **Results** |  |  |
| Participants | 13* | (a) Report numbers of individuals at each stage of study—eg numbers potentially eligible, examined for eligibility, confirmed eligible, included in the study, completing follow-up, and analysed<br>(b) Give reasons for non-participation at each stage<br>(c) Consider use of a flow diagram |
| Descriptive data | 14* | (a) Give characteristics of study participants (eg demographic, clinical, social) and information on exposures and potential confounders<br>(b) Indicate number of participants with missing data for each variable of interest<br>(c) *Cohort study*—Summarise follow-up time (eg, average and total amount) |
| Outcome data | 15* | *Cohort study*—Report numbers of outcome events or summary measures over time<br>*Case-control study*—Report numbers in each exposure category, or summary measures of exposure<br>*Cross-sectional study*—Report numbers of outcome events or summary measures |
| Main results | 16 | (a) Give unadjusted estimates and, if applicable, confounder-adjusted estimates and their precision (eg, 95% confidence interval). Make clear which confounders were adjusted for and why they were included<br>(b) Report category boundaries when continuous variables were categorized<br>(c) If relevant, consider translating estimates of relative risk into absolute risk for a meaningful time period |
| Other analyses | 17 | Report other analyses done—eg analyses of subgroups and interactions, and sensitivity analyses |
| **Discussion** |  |  |
| Key results | 18 | Summarise key results with reference to study objectives |
| Limitations | 19 | Discuss limitations of the study, taking into account sources of potential bias or imprecision. Discuss both direction and magnitude of any potential bias |
| Interpretation | 20 | Give a cautious overall interpretation of results considering objectives, limitations, multiplicity of analyses, results from similar studies, and other relevant evidence |
| Generalisability | 21 | Discuss the generalisability (external validity) of the study results |
| **Other information** |  |  |
| Funding | 22 | Give the source of funding and the role of the funders for the present study and, if applicable, for the original study on which the present article is based |

\*Give information separately for cases and controls in case-control studies and, if applicable, for exposed and unexposed groups in cohort and cross-sectional studies.
**Note:** An Explanation and Elaboration article discusses each checklist item and gives methodological background and published examples of transparent reporting. The STROBE checklist is best used in conjunction with this article (freely available on the Web sites of PLoS Medicine at http://www.plosmedicine.org/, Annals of Internal Medicine at http://www.annals.org/, and Epidemiology at http://www.epidem.com/). Information on the STROBE Initiative is available at www.strobe-statement.org.

---

**eTable 1.** Frequency of ICD-10 diagnoses among younger children aged 0-4 years (n = 45)

| ICD-10 Code | Description | n | % |
| --- | --- | --- | --- |
| J06 | URTI | 19 | 42.2 |
| E34.3 | Short Stature | 6 | 13.3 |
| K59 | Constipation | 6 | 13.3 |
| L30.9 | Eczema | 6 | 13.3 |
| R21 | Rash | 6 | 13.3 |
| H66.9 | Otitis Media | 5 | 11.1 |
| A09.0 | Diarrhea | 3 | 6.7 |
| E61.1 | Iron Deficiency | 3 | 6.7 |
| J35.3 | Enlarged Tonsils and Adenoids | 3 | 6.7 |
| A07.1 | Giardia | 2 | 4.4 |
| D48.5 | Lesion (Skin Lesions) | 2 | 4.4 |
| D50 | Iron Deficiency Anemia | 2 | 4.4 |
| D56.3 | Beta Thalassemia Trait | 2 | 4.4 |
| H50 | Strabismus | 2 | 4.4 |
| J02.9 | Pharyngitis | 2 | 4.4 |
| M79.6 | Pain in Limb | 2 | 4.4 |
| N47.1 | Phimosis | 2 | 4.4 |
| N48.1 | Balantis | 2 | 4.4 |
| Q82.6 | Sacral Dimple | 2 | 4.4 |
| R62.8 | Low Weight | 2 | 4.4 |
| R78.71 | Abnormal Lead Level in Blood | 2 | 4.4 |
| A06 | Entamoeba Histolytica | 1 | 2.2 |
| A07.8 | Dientamoeba Fragilis | 1 | 2.2 |
| A08.4 | Viral Gastroenteritis | 1 | 2.2 |
| B08.1 | Molluscum Contagiosum | 1 | 2.2 |
| B36.0 | Tinea Versicolor | 1 | 2.2 |
| B71.0 | Hymenolepis nana | 1 | 2.2 |
| B78 | Strongyloides | 1 | 2.2 |
| E55 | Vitamin D Deficiency | 1 | 2.2 |
| E66 | Obesity | 1 | 2.2 |
| F80.9 | Speech Delay | 1 | 2.2 |
| H10 | Conjunctivitis | 1 | 2.2 |
| J40 | Bronchitis | 1 | 2.2 |
| J45 | Asthma | 1 | 2.2 |
| K02 | Dental Caries | 1 | 2.2 |
| L08.9 | Skin Infection (Buttock) | 1 | 2.2 |
| L21 | Seborrheic Dermatitis | 1 | 2.2 |
| L29.8 | Genital Pruritus | 1 | 2.2 |
| L81.9 | Hypopigmented Lesions | 1 | 2.2 |
| L85.3 | Dry Skin | 1 | 2.2 |
| M25.5 | Knee Pain | 1 | 2.2 |
| N39.0 | Urinary Tract infection | 1 | 2.2 |
| N50.89 | Scrotal Mass | 1 | 2.2 |
| P12.0 | Cephalhematoma | 1 | 2.2 |
| P59.9 | Neonatal Jaundice | 1 | 2.2 |
| Q53.1 | Undescended testicles | 1 | 2.2 |
| Q55.22 | Retractile Testes | 1 | 2.2 |
| Q67.3 | Plagiocephaly | 1 | 2.2 |
| R04.0 | Epistaxis | 1 | 2.2 |
| R05 | Cough | 1 | 2.2 |
| R06 | SOB | 1 | 2.2 |
| R09.81 | Congestion | 1 | 2.2 |
| R10 | Abdominal Pain | 1 | 2.2 |
| R11 | Vomiting | 1 | 2.2 |
| R56.00 | Febrile Seizures | 1 | 2.2 |
| R59.0 | Lymphadenopathy | 1 | 2.2 |
| R63.0 | Poor Appetite | 1 | 2.2 |
| Z41.2 | Circumcision | 1 | 2.2 |

**eTable 2.** Frequency of ICD-10 diagnoses among older children aged 5-11 years (n=37)

| ICD-10 Code | Description | n | % |
| --- | --- | --- | --- |
| A07.1 | Giardia | 11 | 29.7 |
| R10 | Abdominal Pain | 7 | 18.9 |
| K02 | Dental Caries | 6 | 16.2 |
| E61.1 | Iron Deficiency | 5 | 13.5 |
| E34.3 | Short Stature | 4 | 10.8 |
| J02.9 | Pharyngitis | 4 | 10.8 |
| K59 | Constipation | 3 | 8.1 |
| A07.8 | Dientamoeba Fragilis | 2 | 5.4 |
| B07 | Plantar Wart | 2 | 5.4 |
| B80 | Enterobius Vermicularis | 2 | 5.4 |
| D56.3 | Beta Thalassemia Trait | 2 | 5.4 |
| D72.1 | Eosinophilia | 2 | 5.4 |
| J06 | URTI | 2 | 5.4 |
| J45 | Asthma | 2 | 5.4 |
| L30.5 | Tinea Alba | 2 | 5.4 |
| L30.9 | Eczema | 2 | 5.4 |
| L70 | Acne | 2 | 5.4 |
| L85.3 | Dry Skin | 2 | 5.4 |
| M25.5 | Knee Pain | 2 | 5.4 |
| M79.6 | Pain in Limb | 2 | 5.4 |
| R04.0 | Epistaxis | 2 | 5.4 |
| R05 | Cough | 2 | 5.4 |
| R35 | Polyuria | 2 | 5.4 |
| R50.9 | Fever | 2 | 5.4 |
| R62.8 | Low Weight | 2 | 5.4 |
| A06 | Entamoeba Histolytica | 1 | 2.7 |
| B71.0 | Hymenolepis nana | 1 | 2.7 |
| D50 | Iron Deficiency Anemia | 1 | 2.7 |
| D56.1 | Beta Thalassemia D56.1 | 1 | 2.7 |
| D64 | Anemia | 1 | 2.7 |
| E45 | Stunting | 1 | 2.7 |
| E66 | Obesity | 1 | 2.7 |
| F39 | Mood Disorder (Unspecified) | 1 | 2.7 |
| F51.5 | Nightmares | 1 | 2.7 |
| G25.81 | Restless Leg Syndrome | 1 | 2.7 |
| G43 | Migraine | 1 | 2.7 |
| G44.2 | Tension Headache | 1 | 2.7 |
| H10 | Conjunctivitis | 1 | 2.7 |
| H50.1 | Exotropia | 1 | 2.7 |
| J32.9 | Sinusitis | 1 | 2.7 |
| K09.9 | Oral Cyst | 1 | 2.7 |
| K90.0 | Celiac Disease | 1 | 2.7 |
| L04.0 | Lymphadenopathy | 1 | 2.7 |
| L08.9 | Skin Infection (Buttock) | 1 | 2.7 |
| L29.8 | Genital Pruritus | 1 | 2.7 |
| L81.8 | Hyperpigmented Skin Lesion | 1 | 2.7 |
| L98.9 | Congenital Skin Disorder | 1 | 2.7 |
| M89.28 | Delayed Bone Age | 1 | 2.7 |
| N39.44 | Nocturnal Enuresis | 1 | 2.7 |
| Q16 | Congenital Deafness | 1 | 2.7 |
| Q53.1 | Undescended testicles | 1 | 2.7 |
| Q55.22 | Retractile Testes | 1 | 2.7 |
| R11 | Vomiting | 1 | 2.7 |
| R22.0 | Facial Swelling | 1 | 2.7 |
| R30 | Dysuria R30 | 1 | 2.7 |
| R59.0 | Lymphadenopathy | 1 | 2.7 |
| R63.0 | Poor Appetite | 1 | 2.7 |
| R78.71 | Abnormal Lead Level in Blood | 1 | 2.7 |
| R82.4 | Ketonuria | 1 | 2.7 |

**eTable 3.** Frequency of ICD-10 diagnoses among adolescents aged 12-17 years (n=28)

| ICD-10 Code | Description | n | % |
| --- | --- | --- | --- |
| B98.0 | H Pylori | 7 | 25.0 |
| L70 | Acne | 6 | 21.4 |
| A07.1 | Giardia | 5 | 17.9 |
| M25.5 | Knee Pain | 4 | 14.3 |
| N94.6 | Dysmenorrhea | 4 | 14.3 |
| D50.8 | Microcytosis | 3 | 10.7 |
| D56.3 | Beta Thalassemia Trait | 3 | 10.7 |
| E34.3 | Short Stature | 3 | 10.7 |
| E61.1 | Iron Deficiency | 3 | 10.7 |
| G43 | Migraine | 3 | 10.7 |
| K59 | Constipation | 3 | 10.7 |
| M79.6 | Pain in Limb | 3 | 10.7 |
| R51 | Headache | 3 | 10.7 |
| R62.8 | Low Weight | 3 | 10.7 |
| B07 | Plantar Wart | 2 | 7.1 |
| E53 | Vitamin B12 Deficiency | 2 | 7.1 |
| E55 | Vitamin D Deficiency | 2 | 7.1 |
| E66 | Obesity | 2 | 7.1 |
| H61.2 | Impacted Cerumen | 2 | 7.1 |
| L21.0 | Seborrhea capitis | 2 | 7.1 |
| L65 | Hair Loss | 2 | 7.1 |
| L67.1 | Grey hair | 2 | 7.1 |
| M54.5 | Mechanical Back Pain | 2 | 7.1 |
| R10.9 | Unspecified abdominal pain | 2 | 7.1 |
| A07.2 | Cryptosporidium | 1 | 3.6 |
| B37.3 | Vaginal Candidiasis | 1 | 3.6 |
| B71.0 | Hymenolepis nana | 1 | 3.6 |
| B80 | Enterobius Vermicularis | 1 | 3.6 |
| B85.0 | Head Lice | 1 | 3.6 |
| D72.1 | Eosinophilia | 1 | 3.6 |
| E06.9 | Thyroiditis | 1 | 3.6 |
| E45 | Stunting | 1 | 3.6 |
| F32.9 | Depression | 1 | 3.6 |
| F34.1 | Dysthymia | 1 | 3.6 |
| F41 | Anxiety | 1 | 3.6 |
| F51.0 | Insomnia | 1 | 3.6 |
| F51.5 | Nightmares | 1 | 3.6 |
| F88 | Developmental Disorder | 1 | 3.6 |
| G80.8 | Cerebral Palsy | 1 | 3.6 |
| H11.0 | Pterygium | 1 | 3.6 |
| H73.90 | Abnormal Tympanic Membranes | 1 | 3.6 |
| H91 | Hearing Loss | 1 | 3.6 |
| I95.1 | Orthostatic Hypotension | 1 | 3.6 |
| J02.9 | Pharyngitis | 1 | 3.6 |
| K02 | Dental Caries | 1 | 3.6 |
| K12.0 | Aphthous Ulcer | 1 | 3.6 |
| K21 | GERD | 1 | 3.6 |
| L80 | Vitiligo | 1 | 3.6 |
| L81.2 | Freckles | 1 | 3.6 |
| L81.8 | Hyperpigmented Skin Lesion | 1 | 3.6 |
| L83 | Acanthosis Nigricans | 1 | 3.6 |
| L85.3 | Dry Skin | 1 | 3.6 |
| L85.8 | Keratosis Pilaris | 1 | 3.6 |
| N13.3 | Hydronephrosis | 1 | 3.6 |
| N20.0 | Kidney Stone | 1 | 3.6 |
| O03 | Spontaneous Abortion | 1 | 3.6 |
| R10 | Abdominal Pain | 1 | 3.6 |
| R11 | Vomiting | 1 | 3.6 |
| R16.0 | Hepatomegaly | 1 | 3.6 |
| R16.1 | Splenomegaly. | 1 | 3.6 |
| R21 | Rash | 1 | 3.6 |
| R41.840 | Lack of Concentration | 1 | 3.6 |
| R53 | Fatigue | 1 | 3.6 |
| R56.8 | Other and unspecified convulsions | 1 | 3.6 |

**eTable 4.** Frequency of ICD-10 diagnoses among female adults aged ≥ 18 year (n = 99)

| ICD-10 Code | Description | n | % |
| --- | --- | --- | --- |
| R10 | Abdominal Pain | 38 | 38.4 |
| M54.5 | Mechanical Back Pain | 26 | 26.3 |
| B98.0 | H Pylori | 20 | 20.2 |
| R51 | Headache | 18 | 18.2 |
| E61.1 | Iron Deficiency | 16 | 16.2 |
| E55 | Vitamin D Deficiency | 15 | 15.2 |
| K21 | GERD | 14 | 14.1 |
| E53 | Vitamin B12 Deficiency | 13 | 13.1 |
| L70 | Acne | 13 | 13.1 |
| E03.9 | Hypothyroidism | 11 | 11.1 |
| G43 | Migraine | 11 | 11.1 |
| K59 | Constipation | 11 | 11.1 |
| M25.5 | Knee Pain | 11 | 11.1 |
| M79.6 | Pain in Limb | 11 | 11.1 |
| A07.1 | Giardia | 10 | 10.1 |
| D50 | Iron Deficiency Anemia | 10 | 10.1 |
| F32.9 | Depression | 10 | 10.1 |
| N76.0 | Bacterial Vaginosis | 10 | 10.1 |
| E78 | Dyslipidemia | 9 | 9.1 |
| F51.0 | Insomnia | 9 | 9.1 |
| I10 | Hypertension | 9 | 9.1 |
| G44.2 | Tension Headache | 8 | 8.1 |
| N94.6 | Dysmenorrhea | 8 | 8.1 |
| R21 | Rash | 8 | 8.1 |
| D64 | Anemia | 7 | 7.1 |
| E11 | Diabetes Mellitus Type 2 | 7 | 7.1 |
| R53 | Fatigue | 7 | 7.1 |
| K29.7 | Gastritis | 6 | 6.1 |
| L65 | Hair Loss | 6 | 6.1 |
| M19 | Osteoarthritis | 6 | 6.1 |
| N89.8 | Vaginal Discharge | 6 | 6.1 |
| N94.1 | Dyspareunia | 6 | 6.1 |
| R07 | Chest Pain | 6 | 6.1 |
| A07.8 | Dientamoeba Fragilis | 5 | 5.1 |
| J06 | URTI | 5 | 5.1 |
| L30.9 | Eczema | 5 | 5.1 |
| L81.1 | Melasma | 5 | 5.1 |
| N39.0 | Urinary Tract infection | 5 | 5.1 |
| N92.6 | Irregular Menstruation | 5 | 5.1 |
| R05 | Cough | 5 | 5.1 |
| R42 | Vertigo | 5 | 5.1 |
| R52 | Pain, unspecified | 5 | 5.1 |
| B37.3 | Vaginal Candidiasis | 4 | 4.0 |
| D50.8 | Microcytosis | 4 | 4.0 |
| D72.8 | Lymphocytosis | 4 | 4.0 |
| F41 | Anxiety | 4 | 4.0 |
| K02 | Dental Caries | 4 | 4.0 |
| K64 | Hemorrhoids | 4 | 4.0 |
| N93.9 | Abnormal Uterine Bleeding | 4 | 4.0 |
| A15.9 | Latent tuberculosis | 3 | 3.0 |
| B00.1 | Herpes Simplex | 3 | 3.0 |
| B35.4 | Tinea Corporis | 3 | 3.0 |
| D56.3 | Beta Thalassemia Trait | 3 | 3.0 |
| E04 | Goiter/Goitre | 3 | 3.0 |
| F43.1 | PTSD | 3 | 3.0 |
| F43.2 | Adjustment Disorder | 3 | 3.0 |
| H54 | Blurring of Vision | 3 | 3.0 |
| H61.2 | Impacted Cerumen | 3 | 3.0 |
| I83.9 | Varicose Veins<br>(Superficial) | 3 | 3.0 |
| J02.9 | Pharyngitis | 3 | 3.0 |
| J32.9 | Sinusitis | 3 | 3.0 |
| M06.9 | Rheumatoid arthritis,<br>unspecified | 3 | 3.0 |
| M72.2 | Plantar Fasciitis | 3 | 3.0 |
| M75 | Rotator cuff syndrome | 3 | 3.0 |
| M81.9 | Osteoporosis | 3 | 3.0 |
| N64.4 | Breast Pain | 3 | 3.0 |
| N91.2 | Amenorrhea | 3 | 3.0 |
| N92.0 | Menorrhagia | 3 | 3.0 |
| O03 | Spontaneous Abortion | 3 | 3.0 |
| R00.2 | Palpitations | 3 | 3.0 |
| R25.2 | Cramping | 3 | 3.0 |
| R80 | Proteinuria | 3 | 3.0 |
| B18.2 | Chronic viral hepatitis C | 2 | 2.0 |
| D50.9 | Normocytic Anemia | 2 | 2.0 |
| E05.9 | Hyperthyroidism,<br>unspecified | 2 | 2.0 |
| E66 | Obesity | 2 | 2.0 |
| F43.8 | Stress | 2 | 2.0 |
| H04.129 | Dry eye | 2 | 2.0 |
| H10 | Conjunctivitis | 2 | 2.0 |
| H11.0 | Pterygium | 2 | 2.0 |
| H57.12 | Ocular pain, left eye | 2 | 2.0 |
| H72 | TM perforation | 2 | 2.0 |
| H91 | Hearing Loss | 2 | 2.0 |
| J30.1 | Allergic rhinitis due to<br>pollen | 2 | 2.0 |
| J34.3 | hypertrophy of turbinate | 2 | 2.0 |
| K04.7 | Dental Abscess | 2 | 2.0 |
| L21 | Seborrheic Dermatitis | 2 | 2.0 |
| L29 | Pruritus | 2 | 2.0 |
| L29.3 | Anogenital Pruritus | 2 | 2.0 |
| L85.3 | Dry Skin | 2 | 2.0 |
| M17.0 | Bilateral primary<br>osteoarthritis of knee | 2 | 2.0 |
| M62.6 | Muscle Strain | 2 | 2.0 |
| M75.0 | Adhesive capsulitis of<br>shoulder | 2 | 2.0 |
| M75.5 | Subacromial bursitis | 2 | 2.0 |
| M85.8 | Osteopenia | 2 | 2.0 |
| N91.1 | Secondary Amenorrhea | 2 | 2.0 |
| O03.6 | Inevitable Spontaneous<br>Abortion | 2 | 2.0 |
| R06 | Abnormalities of<br>breathing | 2 | 2.0 |
| R30 | Dysuria | 2 | 2.0 |
| R31 | Hematuria | 2 | 2.0 |
| R41.3 | Memory Loss | 2 | 2.0 |
| R59.0 | Lymphadenopathy | 2 | 2.0 |
| R62.8 | Low Weight | 2 | 2.0 |
| R63.4 | Weight Loss | 2 | 2.0 |
| T79.6 | Shin Splints | 2 | 2.0 |
| Z60.4 | Social Isolation | 2 | 2.0 |
| A06 | Entamoeba Histolytica | 1 | 1.0 |
| A09.0 | Other and unspecified<br>gastroenteritis and colitis<br>of infectious origin | 1 | 1.0 |
| A09.9 | Gastroenteritis and colitis<br>of unspecified origin | 1 | 1.0 |
| A16.7 | Tuberculosis | 1 | 1.0 |
| B02.9 | Zoster NOS | 1 | 1.0 |
| B35.1 | Onychomycosis | 1 | 1.0 |
| B65 | Schistosomiasis | 1 | 1.0 |
| B77.9 | Ascariasis | 1 | 1.0 |
| B78 | Strongyloides | 1 | 1.0 |
| B81.1 | Melasma | 1 | 1.0 |
| B89 | Blasocystis Hominis | 1 | 1.0 |
| B95.1 | GBS Bacteruria | 1 | 1.0 |
| D17.9 | Angiolipoma | 1 | 1.0 |
| D24.9 | Benign Breast Lesion | 1 | 1.0 |
| D25.9 | Uterine Fibroid | 1 | 1.0 |
| D48.9 | Breast Lesion | 1 | 1.0 |
| D56.1 | Beta Thalassemia | 1 | 1.0 |
| D69.6 | Thrombocytopenia | 1 | 1.0 |
| D70 | Neutropenia | 1 | 1.0 |
| D75.8 | Macrocytosis | 1 | 1.0 |
| E06.9 | Thyroiditis | 1 | 1.0 |
| E23.0 | Hypopituitary Disease | 1 | 1.0 |
| E28.2 | PCOS | 1 | 1.0 |
| E28.319 | Premature Menopause | 1 | 1.0 |
| E28.39 | Primary Ovarian<br>Insufficiency | 1 | 1.0 |
| E65 | Abdominal Obesity | 1 | 1.0 |
| E86 | Dehydration | 1 | 1.0 |
| F20.9 | schizophrenia | 1 | 1.0 |
| F25.9 | Schizoaffective Disorder | 1 | 1.0 |
| F34.1 | Dysthymia | 1 | 1.0 |
| F43.8 | Stress | 1 | 1.0 |
| F45.8 | Globus Sensation | 1 | 1.0 |
| F52.0 | Low Libido | 1 | 1.0 |
| F53.0 | Post-Partum Depression | 1 | 1.0 |
| F79 | Intellectual Disability | 1 | 1.0 |
| G25.81 | Restless Leg Syndrome | 1 | 1.0 |
| G40 | Epilepsy | 1 | 1.0 |
| G57.1 | Meralgia Paresthetica | 1 | 1.0 |
| H00.01 | Hordeolum externum | 1 | 1.0 |
| H40 | Glaucoma | 1 | 1.0 |
| H50 | Strabismus | 1 | 1.0 |
| H57.1 | Eye Pain | 1 | 1.0 |
| H60.9 | Unspecified otitis externa | 1 | 1.0 |
| H66.9 | Otitis Media | 1 | 1.0 |
| H69.82 | Other specified disorders<br>of Eustachian tube, left ear | 1 | 1.0 |
| H74.0 | Tympanosclerosis | 1 | 1.0 |
| H92.0 | Ear Pain | 1 | 1.0 |
| I09.9 | Rheumatic heart disease | 1 | 1.0 |
| I26 | Pulmonary Embolism | 1 | 1.0 |
| I85 | Esophageal Varices | 1 | 1.0 |
| J34.89 | Dry Nose | 1 | 1.0 |
| J45 | Asthma | 1 | 1.0 |
| J47 | Bronchiectasis | 1 | 1.0 |
| J84.1 | Interstitial Fibrosis | 1 | 1.0 |
| K08.8 | Tooth Pain | 1 | 1.0 |
| K12.0 | Aphthous Ulcer | 1 | 1.0 |
| K45 | Abdominal Hernia | 1 | 1.0 |
| K58 | Irritable Bowel Syndrome | 1 | 1.0 |
| K60.2 | Anal Fissure | 1 | 1.0 |
| K74.6 | Other and unspecified<br>cirrhosis of liver | 1 | 1.0 |
| K76.0 | Fatty Liver | 1 | 1.0 |
| K76.9 | Liver Mass | 1 | 1.0 |
| K80 | Cholelithiasis | 1 | 1.0 |
| K80.5 | Biliary Colic | 1 | 1.0 |
| K81.0 | Acute Cholecystitis | 1 | 1.0 |
| L24.9 | Irritant Dermatitis | 1 | 1.0 |
| L30.1 | Dyshydrotic Eczema | 1 | 1.0 |
| L50 | Urticaria | 1 | 1.0 |
| L68.0 | Hirsutism | 1 | 1.0 |
| L81.2 | Freckles | 1 | 1.0 |
| L94.0 | Scleroderma | 1 | 1.0 |
| M13.9 | Inflammatory Arthritis | 1 | 1.0 |
| M22.2 | Patellofemoral Disorders | 1 | 1.0 |
| M25.561 | Pain in right knee | 1 | 1.0 |
| M26.62 | Arthralgia | 1 | 1.0 |
| M34.9 | Systemic Sclerosis | 1 | 1.0 |
| M41.9 | Scoliosis | 1 | 1.0 |
| M54.1 | Lumbar Radiculopathy | 1 | 1.0 |
| M54.2 | Neck Pain | 1 | 1.0 |
| M54.3 | Sciatica | 1 | 1.0 |
| M75.41 | Impingement syndrome of right shoulder | 1 | 1.0 |
| M75.81 | Other should lesions, right shoulder | 1 | 1.0 |
| M76.2 | Iliotibial band syndrome | 1 | 1.0 |
| M79.0 | Rheumatism | 1 | 1.0 |
| M94.0 | Costochondritis | 1 | 1.0 |
| N10 | Acute Pyelonephritis | 1 | 1.0 |
| N13.3 | Hydronephrosis | 1 | 1.0 |
| N18.9 | Chronic Renal Failure | 1 | 1.0 |
| N20.0 | Kidney Stone | 1 | 1.0 |
| N63 | Breast Nodule | 1 | 1.0 |
| N64.59 | Breast Itch | 1 | 1.0 |
| N80 | Adenomyosis | 1 | 1.0 |
| N85.0 | Endometrial Hyperplasia | 1 | 1.0 |
| N88.8 | Nabothian Cyst | 1 | 1.0 |
| N91.5 | Oligomenorrhea | 1 | 1.0 |
| N92.1 | Metrorrhagia | 1 | 1.0 |
| N94.2 | Vaginismus | 1 | 1.0 |
| N94.3 | Premenstrual Tension Syndrome | 1 | 1.0 |
| N95.0 | Postmenopausal Bleeding | 1 | 1.0 |
| N97.9 | Female infertility, unspecified | 1 | 1.0 |
| O85 | Post-Partum Endometritis | 1 | 1.0 |
| P05.1 | Small for Gestational Age | 1 | 1.0 |
| Q21.2 | Atrioventricular septal defect | 1 | 1.0 |
| Q68.4 | Congenital Bowing of Tibial Bones | 1 | 1.0 |
| R01.1 | Cardiac murmur, unspecified | 1 | 1.0 |
| R07.0 | Sore Throat | 1 | 1.0 |
| R11 | Vomiting | 1 | 1.0 |
| R14 | Bloating | 1 | 1.0 |
| R16.1 | Splenomegaly | 1 | 1.0 |
| R19.00 | Pelvic Mass | 1 | 1.0 |
| R19.06 | Renal Mass | 1 | 1.0 |
| R19.6 | Halitosis | 1 | 1.0 |
| R20.8 | Paresthesia | 1 | 1.0 |
| R22.1 | Neck Mass | 1 | 1.0 |
| R41.840 | Lack of Concentration | 1 | 1.0 |
| R45.8 | Suicidal Ideation | 1 | 1.0 |
| R55 | Syncope | 1 | 1.0 |
| R73.0 | Prediabetes | 1 | 1.0 |
| S13.6 | Neck strain | 1 | 1.0 |
| S39 | Other and unspecified injuries of the abdomen, back, pelvis, and external genitalia | 1 | 1.0 |
| S60.00 | Finger Contusion | 1 | 1.0 |
| T14.0 | Infected Blister Finger | 1 | 1.0 |
| T75.3 | Motion Sickness | 1 | 1.0 |
| U00 | COVID-19 | 1 | 1.0 |
| Z85.3 | History of Breast Cancer | 1 | 1.0 |

**eTable 5.** Frequency of ICD-10 diagnoses among male adults aged ≥ 18 year (n = 89)

| ICD-10 Code | Description | n | % |
| --- | --- | --- | --- |
| B98.0 | H Pylori | 24 | 27.0 |
| R10 | Abdominal Pain | 22 | 24.7 |
| M54.5 | Mechanical Back Pain | 20 | 22.5 |
| M25.5 | Knee Pain | 16 | 18.0 |
| E55 | Vitamin D Deficiency | 14 | 15.7 |
| E78 | Dyslipidemia | 14 | 15.7 |
| I10 | Hypertension | 11 | 12.4 |
| K21 | Gastroesophageal reflux disease | 9 | 10.1 |
| K59 | Constipation | 9 | 10.1 |
| A07.1 | Giardia | 7 | 7.9 |
| F41 | Anxiety | 7 | 7.9 |
| K29.7 | Gastritis | 7 | 7.9 |
| R07 | Chest Pain | 7 | 7.9 |
| R51 | Headache | 7 | 7.9 |
| H91 | Hearing Loss | 6 | 6.7 |
| J06 | URTI | 6 | 6.7 |
| K02 | Dental Caries | 6 | 6.7 |
| M79.6 | Pain in Limb | 6 | 6.7 |
| A07.8 | Dientamoeba Fragilis | 5 | 5.6 |
| F32.9 | Depression | 5 | 5.6 |
| J02.9 | Pharyngitis | 5 | 5.6 |
| L65 | Hair Loss | 5 | 5.6 |
| L70 | Acne | 5 | 5.6 |
| A15.9 | Latent TB | 4 | 4.5 |
| D56.3 | Beta Thalassemia Trait | 4 | 4.5 |
| D70 | Neutropenia | 4 | 4.5 |
| E11 | Diabetes Mellitus Type 2 | 4 | 4.5 |
| F43.1 | PTSD | 4 | 4.5 |
| F43.8 | Stress | 4 | 4.5 |
| F51.0 | Insomnia | 4 | 4.5 |
| R21 | Rash | 4 | 4.5 |
| R42 | Vertigo | 4 | 4.5 |
| R52 | Pain, unspecified | 4 | 4.5 |
| D17.9 | Angiolipoma | 3 | 3.4 |
| D50.8 | Microcytosis | 3 | 3.4 |
| E53 | Vitamin B12 Deficiency | 3 | 3.4 |
| E61.1 | Iron Deficiency | 3 | 3.4 |
| E66 | Obesity | 3 | 3.4 |
| F43.2 | Adjustment Disorder | 3 | 3.4 |
| H53 | Visual Problem | 3 | 3.4 |
| H61.2 | Impacted Cerumen | 3 | 3.4 |
| L85.3 | Dry Skin | 3 | 3.4 |
| M79.1 | Myalgia | 3 | 3.4 |
| R05 | Cough | 3 | 3.4 |
| R09.81 | Congestion | 3 | 3.4 |
| A09.0 | Diarrhea | 2 | 2.2 |
| B16 | Hepatitis B | 2 | 2.2 |
| B18.1 | Chronic Hepatitis B | 2 | 2.2 |
| D50 | Iron Deficiency Anemia | 2 | 2.2 |
| D69.6 | Thrombocytopenia | 2 | 2.2 |
| E03.9 | Hypothyroidism | 2 | 2.2 |
| E05.9 | Hyperthyroidism | 2 | 2.2 |
| E65 | Abdominal Obesity | 2 | 2.2 |
| G43 | Migraine | 2 | 2.2 |
| G62.9 | Polyneuropathy,<br>unspecified | 2 | 2.2 |
| H52 | Disorders of Refraction | 2 | 2.2 |
| H54 | Blurring of Vision | 2 | 2.2 |
| I64 | Cerebrovascular Accident | 2 | 2.2 |
| J30 | rhinitis | 2 | 2.2 |
| K12.0 | Aphthous Ulcer | 2 | 2.2 |
| K40 | Inguinal Hernia | 2 | 2.2 |
| K58 | Irritable Bowel Syndrome | 2 | 2.2 |
| K64 | Hemorrhoids | 2 | 2.2 |
| L29 | Pruritus | 2 | 2.2 |
| L30.9 | Eczema | 2 | 2.2 |
| L81.8 | Hyperpigmented Skin Lesion | 2 | 2.2 |
| L90.5 | Scar | 2 | 2.2 |
| M19 | Osteoarthritis | 2 | 2.2 |
| N28.1 | Kidney Cyst | 2 | 2.2 |
| Q82 | Skin Tags | 2 | 2.2 |
| R11 | Vomiting | 2 | 2.2 |
| R22.1 | Neck Mass | 2 | 2.2 |
| R53 | Fatigue | 2 | 2.2 |
| S63.5 | Other and unspecified sprain of wrist | 2 | 2.2 |
| S93.4 | Ankle Sprain | 2 | 2.2 |
| A09.9 | Diarrhea | 1 | 1.1 |
| A53.0 | Latent Syphilis | 1 | 1.1 |
| A56.2 | Chlamydia | 1 | 1.1 |
| B00.1 | Herpes Simplex | 1 | 1.1 |
| B07 | Plantar Wart | 1 | 1.1 |
| B35.1 | Onychomycosis | 1 | 1.1 |
| B35.3 | Tinea Pedis | 1 | 1.1 |
| B55.1 | Cutaneous Leishmaniasis | 1 | 1.1 |
| B64 | Non-pathogenic Protozoa | 1 | 1.1 |
| B65 | Schistosomiasis | 1 | 1.1 |
| B78 | Strongyloidiasis | 1 | 1.1 |
| B85.0 | Head Lice | 1 | 1.1 |
| D50.9 | Normocytic Anemia | 1 | 1.1 |
| D55.0 | G6PD Deficiency | 1 | 1.1 |
| D56.1 | Beta Thalassemia | 1 | 1.1 |
| D57.3 | Sickle Cell Trait | 1 | 1.1 |
| D64 | Anemia | 1 | 1.1 |
| D72.8 | Lymphocytosis | 1 | 1.1 |
| D75.8 | Macrocytosis | 1 | 1.1 |
| E04 | Goiter/Goitre | 1 | 1.1 |
| E05.0 | Graves Disease | 1 | 1.1 |
| E78.0 | Hypercholesterolemia | 1 | 1.1 |
| E87.0 | Hypernatremia | 1 | 1.1 |
| F34.1 | Dysthymia | 1 | 1.1 |
| F40.2 | Specific Phobia | 1 | 1.1 |
| F41.1 | Generalized Anxiety Disorder | 1 | 1.1 |
| F44.5 | Psychogenic Non Epileptic Seizure | 1 | 1.1 |
| F51.5 | Nightmares | 1 | 1.1 |
| F52.2 | Erectile Dysfunction | 1 | 1.1 |
| F52.4 | Premature Ejaculation | 1 | 1.1 |
| G56.3 | Radial Nerve Palsy | 1 | 1.1 |
| G57.1 | Meralgia Paresthetica | 1 | 1.1 |
| G60.9 | Peripheral Neuropathy | 1 | 1.1 |
| H10 | Conjunctivitis | 1 | 1.1 |
| H10.1 | Allergic Conjunctivitis | 1 | 1.1 |
| H11.0 | Pterygium | 1 | 1.1 |
| H40 | Glaucoma | 1 | 1.1 |
| H49.11 | Right Fourth Cranial Nerve Palsy | 1 | 1.1 |
| H50 | Strabismus | 1 | 1.1 |
| H57.1 | Eye Pain | 1 | 1.1 |
| H57.8 | Eye Redness | 1 | 1.1 |
| H65 | Serous Otitis Media | 1 | 1.1 |
| H72 | Tympanic Membrane perforation | 1 | 1.1 |
| H92.0 | Ear Pain | 1 | 1.1 |
| H93.1 | Tinnitus | 1 | 1.1 |
| I20 | Stable Angina | 1 | 1.1 |
| I20.0 | Unstable Angina | 1 | 1.1 |
| I34.0 | Mitral Valve Regurgitation | 1 | 1.1 |
| I48 | Atrial fibrillation | 1 | 1.1 |
| I51.7 | Left Ventricular Hypertrophy | 1 | 1.1 |
| I83.9 | Varicose Veins<br>(Superficial) | 1 | 1.1 |
| J03.9 | Tonsillitis | 1 | 1.1 |
| J18.9 | Pneumonia | 1 | 1.1 |
| J32.9 | Sinusitis | 1 | 1.1 |
| J40 | Bronchitis | 1 | 1.1 |
| J44 | COPD | 1 | 1.1 |
| K08.8 | Tooth Pain | 1 | 1.1 |
| K42 | Umbilical Hernia | 1 | 1.1 |
| K62.5 | Rectal Bleeding | 1 | 1.1 |
| L02.9 | Boil | 1 | 1.1 |
| L08.9 | Skin Infection (Buttock) | 1 | 1.1 |
| L29.9 | Leg Pruritus | 1 | 1.1 |
| L60.0 | Ingrown Toenail | 1 | 1.1 |
| L60.3 | Nail Dystrophy | 1 | 1.1 |
| L63 | Alopecia Areata | 1 | 1.1 |
| M25.362 | Left Knee Instability | 1 | 1.1 |
| M25.9 | Facet Joint Disease | 1 | 1.1 |
| M54.1 | Lumbar Radiculopathy | 1 | 1.1 |
| M60 | Myositis | 1 | 1.1 |
| M62.6 | Muscle Strain | 1 | 1.1 |
| M62.8 | Leg weakness | 1 | 1.1 |
| M70.51 | Pes Anserine Bursitis (Left) | 1 | 1.1 |
| M72.2 | Plantar Fasciitis | 1 | 1.1 |
| M75 | Rotator cuff syndrome | 1 | 1.1 |
| M76.5 | Patellar Tendinopathy | 1 | 1.1 |
| M77.00 | Medial Epicondylitis | 1 | 1.1 |
| M77.1 | Lateral Epicondylitis | 1 | 1.1 |
| M81.9 | Osteoporosis | 1 | 1.1 |
| N39.0 | Urinary Tract infection | 1 | 1.1 |
| N39.4 | Urinary Incontinence | 1 | 1.1 |
| N40 | Prostatism | 1 | 1.1 |
| N41 | Prostatitis | 1 | 1.1 |
| N43.3 | Hydrocele | 1 | 1.1 |
| N50.8 | Scrotal Swelling | 1 | 1.1 |
| Q66 | Bilateral Foot Deformity | 1 | 1.1 |
| R00.1 | bradycardia | 1 | 1.1 |
| R00.2 | Palpitations | 1 | 1.1 |
| R04.0 | Epistaxis | 1 | 1.1 |
| R06 | SOB | 1 | 1.1 |
| R07.0 | Sore Throat | 1 | 1.1 |
| R09.82 | Post Nasal Drip | 1 | 1.1 |
| R13 | Dysphagia | 1 | 1.1 |
| R14 | Bloating | 1 | 1.1 |
| R20.8 | Paresthesia | 1 | 1.1 |
| R22.2 | Supraclavicular Mass | 1 | 1.1 |
| R23.2 | Flushing | 1 | 1.1 |
| R30 | Dysuria | 1 | 1.1 |
| R35 | Polyuria | 1 | 1.1 |
| R36 | Penile Discharge | 1 | 1.1 |
| R39.9 | Lower Urinary Tract<br>Symptoms | 1 | 1.1 |
| R41 | Cognitive Decline NOS | 1 | 1.1 |
| R41.3 | Memory Loss | 1 | 1.1 |
| R41.840 | Lack of Concentration | 1 | 1.1 |
| R45.8 | Suicidal Ideation | 1 | 1.1 |
| R50.8 | Fever with chills | 1 | 1.1 |
| R53.1 | Body Weakness | 1 | 1.1 |
| R55 | Syncope | 1 | 1.1 |
| R63.0 | Poor Appetite | 1 | 1.1 |
| R63.4 | Weight Loss | 1 | 1.1 |
| R80 | Proteinuria R80 | 1 | 1.1 |
| R90.0 | Intracranial Lesion | 1 | 1.1 |
| S30.0 | Contusion Back | 1 | 1.1 |
| S51.9 | Laceration Forearm | 1 | 1.1 |
| S53.4 | Elbow-Flexor Tendonitis | 1 | 1.1 |
| S60.0 | Contusion Finger | 1 | 1.1 |
| S67.190A | Crushing Injury Index<br>Finger | 1 | 1.1 |
| S67.31XA | Crushing Injury Right Wrist | 1 | 1.1 |
| S76.811A | Strained Groin | 1 | 1.1 |
| S83.206A | Right Knee Meniscus Tear | 1 | 1.1 |
| T23.201A | Second Degree Burn of<br>Right Hand | 1 | 1.1 |
| T78.4 | Allergies | 1 | 1.1 |
| W57 | Insect Bites | 1 | 1.1 |
| Y07 | Torture | 1 | 1.1 |
| Z74.09 | Other Reduced Mobility | 1 | 1.1 |

**eTable 6.** Annual clinic appointment no-show rate among Afghan refugee cohort.

| Year | Number of clinic appointments | Number of no-show appointments | No-show rate (%) |
| --- | --- | --- | --- |
| 2011 | 340 | 48 | 14.12 |
| 2012 | 748 | 55 | 7.35 |
| 2013 | 987 | 160 | 16.21 |
| 2014 | 453 | 51 | 11.26 |
| 2015 | 337 | 39 | 11.57 |
| 2016 | 160 | 28 | 17.50 |
| 2017 | 202 | 22 | 10.89 |
| 2018 | 223 | 37 | 16.59 |
| 2019 | 744 | 73 | 9.81 |
| 2020 | 841 | 48 | 5.71 |
| Total | 5035 | 561 | 11.14 |

